# Effectiveness of Mass Vaccination in Brazil against Severe COVID-19 Cases

**DOI:** 10.1101/2021.09.10.21263084

**Authors:** Daniel A.M. Villela, Tatiana Guimarães de Noronha, Leonardo S. Bastos, Antonio G. Pacheco, Oswaldo G Cruz, Luiz Max Carvalho, Claudia Torres Codeço, Marcelo Ferreira da Costa Gomes, Flávio Codeço Coelho, Laís Picinini Freitas, Raquel Martins Lana, Victor Bertollo Gomes Porto, Luiz Antônio Bastos Camacho, Claudio José Struchiner

## Abstract

**Background:** Mass vaccination campaigns started in Brazil on January/2021 with CoronaVac followed by ChAdOx1 nCov-19, and BNT162b2 mRNA vaccines. Target populations initially included vulnerable groups such as people older than 80 years, with comorbidities, of indigenous origin, and healthcare workers. Younger age groups were gradually included.

**Methods:** A national cohort of 66.3 million records was compiled by linking registry-certified COVID-19 vaccination records from the Brazilian National Immunization Program with information on severe COVID-19 cases and deaths. Cases and deaths were aggregated by state and age group. Mixed-effects Poisson models were used to estimate the rate of severe cases and deaths among vaccinated and unvaccinated individuals, and the corresponding estimates of vaccine effectiveness by vaccine platform and age group. The study period is from mid-January to mid-July 2021.

**Results:** Estimates of vaccine effectiveness preventing deaths were highest at 97.9% (95% CrI: 93.5-99.8) among 20-39 years old with ChAdOx1 nCov-19, at 82.7% (95% CrI: 80.7-84.6) among 40-59 years old with CoronaVac, and at 89.9% (87.8--91.8) among 40-59 years old with partial immunization of BNT162b2. For all vaccines combined in the full regimen, the effectiveness preventing severe cases among individuals aged 80+ years old was 35.9% (95% CrI: 34.9-36.9) which is lower than that observed for individuals aged 60-79 years (61.0%, 95% CrI: 60.5-61.5).

**Conclusion:** Despite varying effectiveness estimates, Brazil’s population benefited from vaccination in preventing severe COVID-19 outcomes. Results, however, suggest significant vaccine-specific reductions in effectiveness by age, given by differences between age groups 60-79 years and over 80 years.

## Introduction

Brazil started its national COVID-19 vaccination campaign on January 17, 2021, right after exceeding 200,000 confirmed deaths. Since early in the COVID-19 pandemic, higher risks for severe COVID-19 disease and deaths were strongly related to age factors, pointed out by age-skewed distribution of cases and comorbidities (1,2). Due to limitations in vaccine supplies during the first months of the campaign, a prioritization schedule was developed based on these risk evaluations by technical discussions at the National Immunization Program and the WHO SAGE recommendations (3). Priority was given to those at higher risk of severe disease (elderly, chronic health conditions, and disabled population), vulnerable populations, health care workers, and lastly, essential workers. After completion of the aforementioned groups, vaccination was extended to the entire population in sequential order given by decreasing age.

The nationwide vaccination campaign started with two main vaccines, CoronaVac (SinoVac) and ChAdOx1 nCov-19 (AstraZeneca/Oxford University), initially imported to cover demands, and later produced by Instituto Butantan, São Paulo, and Fundação Oswaldo Cruz, Rio de Janeiro, respectively. Full-regimen vaccination (two doses) was completed after a 28-day (CoronaVac) or 12-week (ChAdOx1 nCov-19) recommended intervals. This time difference made the initial vaccinated groups complete the full regimen as early as February (CoronaVac) or mid-April (ChAdOx1 nCov-19). The campaign policy was to provide vaccination doses over time to the states following a distribution proportional to the states’ populations. On April 29, BNT162b2 mRNA vaccine (Pfizer-BioNTech) was integrated into the regular immunization distribution. Later, on June 15, Ad26.COV2.S vaccines (Janssen) were also imported and integrated into the national immunization program. Brazil adopted a 12-week interval for the BNT162b2 vaccines, whereas Ad26.COV2.S required a single dose. COVID-19 vaccine doses have been administered solely by the public health system and recorded in an electronic database managed by the Ministry of Health.

As Brazil started to implement the vaccination in January 2021, the gamma variant of concern (VOC) was already present in a few states, appearing first in the North and quickly disseminating in the country (4). A mix of factors including the emergence of the new variant, a slow pace in advancing vaccination coverage, and easing or absence of restrictions in many cities and states led to another escalation in the number of cases and, consequently, hospitalizations and deaths. Subsequently, the number of reported cases started decreasing nationwide, even though the epidemiological scenario varied across several states.

Here we report on a massive data analysis to assess the Brazilian vaccination program effectiveness over the initial six months, based on a cohort of health records derived from linkage of two national databases: (1) records of vaccination events and (2) severe COVID-19 cases, including deaths. Therefore, an evaluation of the vaccination program as a whole was possible, as well as separate analyzes investigating the rates of hospitalization and deaths among vaccinated individuals vaccinated with ChAdOx1 nCov-19, CoronaVac, and BNT162b2 mRNA vaccines. The statistical analysis involved a mixed-effects model to estimate the rate ratios between vaccinated and unvaccinated individuals in multiple combinations of outcomes, vaccination regimen, and vaccine, adjusting for age groups and state of residency. The evidences from this work were presented to the technical committee for immunization in the Ministry of Health overseeing the COVID-19 vaccination campaign.

## Methods

### Study population

The National Immunization Program (NIP) provided an anonymized dataset containing individual-level data on vaccination and the occurrence of Severe Acute Respiratory Illness (SARI). The dataset was constructed by the NIP through probabilistic linkage of vaccination data registered at the National Network of Health Data (RNDS) and the data related to SARI cases from the Influenza Epidemiological Surveillance System (SIVEP-Gripe), which include hospitalized cases and deaths (details in Supplementary Text). SARI patient’s nasopharyngeal samples are routinely screened for virus detection. Since the emergence of SARS-CoV-2 virus in Brazil in 2020, SARI cases have occurred predominantly due to SARS-CoV-2 infections, accounting for over 98% of the laboratory-confirmed cases of SARI. Cases with a diagnosis of infection by other viruses were excluded. Records with different vaccines in the first and second doses were disregarded, as well as those with absence of first dose information (date or vaccine manufacturer) but with second dose information. The exploratory analysis included all ages. The statistical analysis included only individuals over 20 years of age. The vaccination database included data up to June 30, 2021, whereas data from SIVEP had notifications as late as July 18, 2021.

Population data by age groups and states were extracted from a national projection maintained by the Brazilian Ministry of Health (source: DataSUS).

### Immunization status

The COVID-19 vaccination campaign started on January 17, 2021, the first epoch for the study (T0). Effectiveness was evaluated in groups either partially or fully immunized (≥14 days after the first dose) and fully immunized (≥14 days after the second dose). Effectiveness were considered in the overall vaccination program (all vaccines) as well as vaccine-specific, CoronaVac, ChAdOx1 nCov-19, and BNT162b2. ChAdOx1 nCov-19 (AZD1222), by Oxford/AstraZeneca, was first imported as Covishield and later produced as Vaxzevria. Vaccination effectiveness for BNT162b2 after two-dose regimen were not evaluated due to short time for observing outcomes in the study period. Vaccination with Janssen started later and, by the last day of observation in the cohort, there were significantly fewer individuals and most importantly, very little time to observe the outcomes of interest. The population data permitted an estimation of the unvaccinated population by age group and states

### Case definition - outcomes of interest

Outcomes of interest were serious illness, i.e. confirmed and probable COVID-19 cases with symptoms leading to hospitalization and COVID-19-related deaths, independent of prior hospitalization. The former could also include death subsequently. The outcomes are signaled in the database by the date of symptoms onset and the final case status (death, recovered or ignored). For each person in the cohort, an outcome of serious COVID-19 illness occurred whenever this person was notified as a COVID-19 SARI case in the national database, using the date of symptoms onset as reference. Therefore, a severe COVID-19 illness after immunization, either partially or fully, characterized an immunized case. Cases without a registry of vaccination or with symptoms before vaccination status were classified as unvaccinated cases. For the outcome of death-only, individuals were counted, if death was registered in the electronic record, and their vaccination status was also dependent on the date of immunization and date of symptoms onset. Vaccinated individuals without any registry of severe COVID-19 during the study period were considered as not presenting the outcomes of interest.

### Statistical analysis

Cases of severe COVID-19 were aggregated by age group (20 - 39, 40 - 59, 60 - 79, and 80+ years old), vaccination status (immunized/not immunized), and the states of residency. The analysis involved a mixed-effects Poisson model to evaluate the case rate ratios (RR), and estimates of vaccination effectiveness 1-RR, which were evaluated per age-group, or per targeted population. We also evaluated overall and vaccine-specific effectiveness. The same framework applies for evaluating effectiveness in preventing deaths. Details about the model are given in the Supplementary Text.

**Figure 1:**
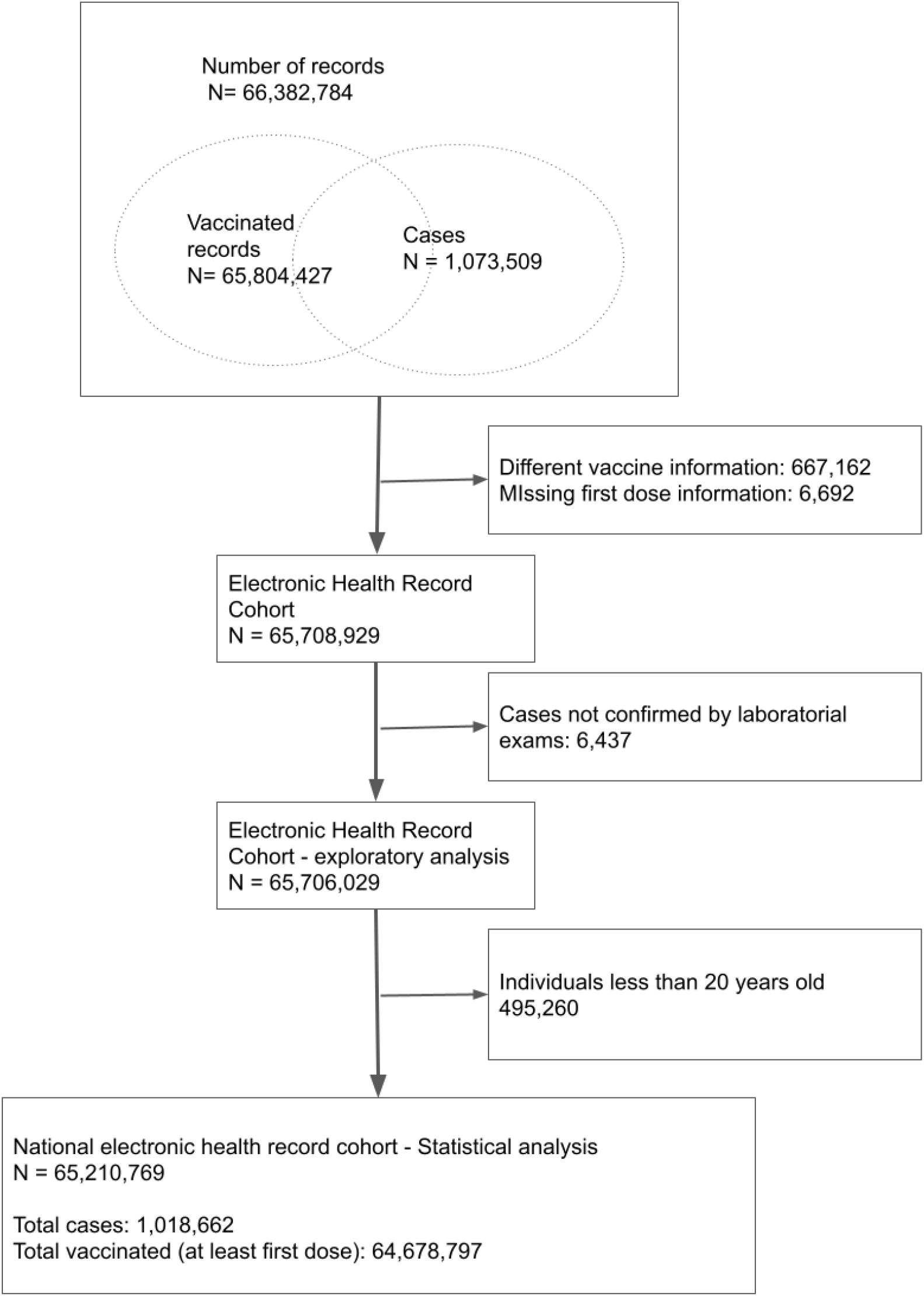
The electronic health record cohort and the databases for exploratory and statistical analysis. The cohort includes both cases from the Brazilian surveillance system (SIVEP-gripe) and the national vaccination databases. After exclusion criteria, there were 784,943 (77%) confirmed cases, and 233,719 (23%) probable cases.

## Results

In the first six months, the majority of first doses (36.6 million) were ChAdOx1 nCov-19 vaccines, whereas most individuals completing full vaccination regimen received CoronaVac (17.3 million) (Table 1). Of individuals with CoronaVac complete regimen, most were in the age groups of 60-79 (65.4%) and 80+ years old (12.0%). Conversely, most cases of fully-immunized individuals with BNT162b2 were in the 20-39 and 40-59 years old groups (33.8% and 61.5%, respectively). For ChAdOx1 nCov-19, the group of 40-59 years old more frequently received at least the first dose (52.5%), whereas individuals fully vaccinated were more concentrated in the age groups of 60-79 (30.4%) and 80+ years old (33.0%). As expected, the Southeast region concentrated most vaccinated individuals due to population sizes, whereas North and Center-West regions had proportionally fewer vaccinations.

**Table 1.** Number of vaccinated individuals by vaccine and regimen (partial to full, and full) in Brazil from January 17 to June 30, 2021. Strata include age groups, sex, and the five regions of Brazil. Numbers for 1+ dose present the number of individuals in the cohort that received at least the first dose, hence also counting the fully vaccinated individuals. Values in percentages indicate the proportions of each stratum item by the total in the stratum/vaccine/regimen.

|  |  | ChAdOx1 nCov-19 |  | CoronaVac |  | BNT162b2 |  |
| --- | --- | --- | --- | --- | --- | --- | --- |
|  |  | 1+ dose (%) | full regimen (%) | 1+ dose (%) | full regimen (%) | 1+ dose (%) | full regimen (%) |
| Age group | 0-19 | 285,064 (0.8) | 15,160 (0.5) | 86,507 ( 0.4) | 53,959 ( 0.3) | 86,244 ( 1.3) | 458 ( 1.2) |
|  | 20-39 | 6,775,915 (18.5) | 586,273 (18.8) | 2,643,137 (12.3) | 1,995,119 (11.5) | 1,806,439 (26.5) | 13,096 (33.8) |
|  | 40-59 | 19,175,389 (52.5) | 539,041 (17.3) | 3,457,567 (16.1) | 1,863,854 (10.8) | 4,749,791 (69.7) | 23,814 (61.5) |
|  | 60-79 | 8,930,724 (24.4) | 945,316 (30.4) | 12,914,363 (60.3) | 11,322,374 (65.4) | 166,351 (2.4) | 1,331 (3.4) |
|  | 80+ | 1,391,144 (3.8) | 1,026,239 (33.0) | 2,319,469 (10.8) | 2,086,627 (12.0) | 3,936 (0.1) | 46 (0.1) |
| Region | North | 2,727,350 ( 7.5) | 394,193 (12.7) | 1,348,827 (6.3) | 1,123,269 (6.5) | 465,931 (6.8) | 77 ( 0.2) |
|  | North east | 8,658,260 (23.7) | 727,709 (23.4) | 4,999,917 (23.3) | 4,166,636 (24.1) | 1,537,377 (22.6) | 285 ( 0.7) |
|  | South east | 16,228,228 (44.4) | 1,317,335 (42.3) | 10,008,792 (46.7) | 7,671,701 (44.3) | 2,983,870 (43.8) | 1,298 (3.4) |
|  | South | 6,096,419 (16.7) | 415,226 (13.3) | 3,479,160 (16.2) | 3,019,037 (17.4) | 1,220,697 (17.9) | 36,901 (95.2) |
|  | Center-West | 2,847,979 (7.8) | 257,566 (8.3) | 1,584,346 (7.4) | 1,341,289 (7.7) | 604,886 (8.9) | 184 (0.5) |
| Sex | F | 20,054,648 (54.9) | 1,926,966 (61.9) | 12,632,426 (59.0) | 10,293,819 (59.4) | 3,901,877 (57.3) | 26,180 (67.6) |
|  | M | 16,503,569<br>(45.1) | 1,185,062<br>(38.1) | 8,788,596<br>(41.0) | 7,028,102<br>(40.6) | 2,910,882<br>(42.7) | 12,565<br>(32.4) |
| Total |  | N = 36,558,236 | N =<br>3,112,029 | N =<br>21,421,043 | N =<br>17,321,933 | N =<br>6,812,761 | N = 38,745 |

Most cases of severe COVID-19 illness (Table 2) among individuals immunized with at least a first dose of ChAdOx1 nCov-19 vaccines were in the age groups of 60-79 and 80+ years old (40.7% and 35.4%, respectively). Of cases in fully immunized individuals with ChAdOx1 nCov-19, 74.6% were 80+ years or older. Most hospitalized cases among CoronaVac immunized individuals occurred in the age group of 60-79 years old, 59.5% of them with at least one dose and 53.7% with two doses. Hospitalized cases among BNT162b2-immunized (partially) individuals were concentrated in the age group of 40-59 years old. Deaths among fully-immunized individuals were 80.5% (ChAdOx1 nCov-19 group) and exceeded 90% counting both groups 60-79 and 80+ receiving CoronaVac (Table 3).

**Table 2:**
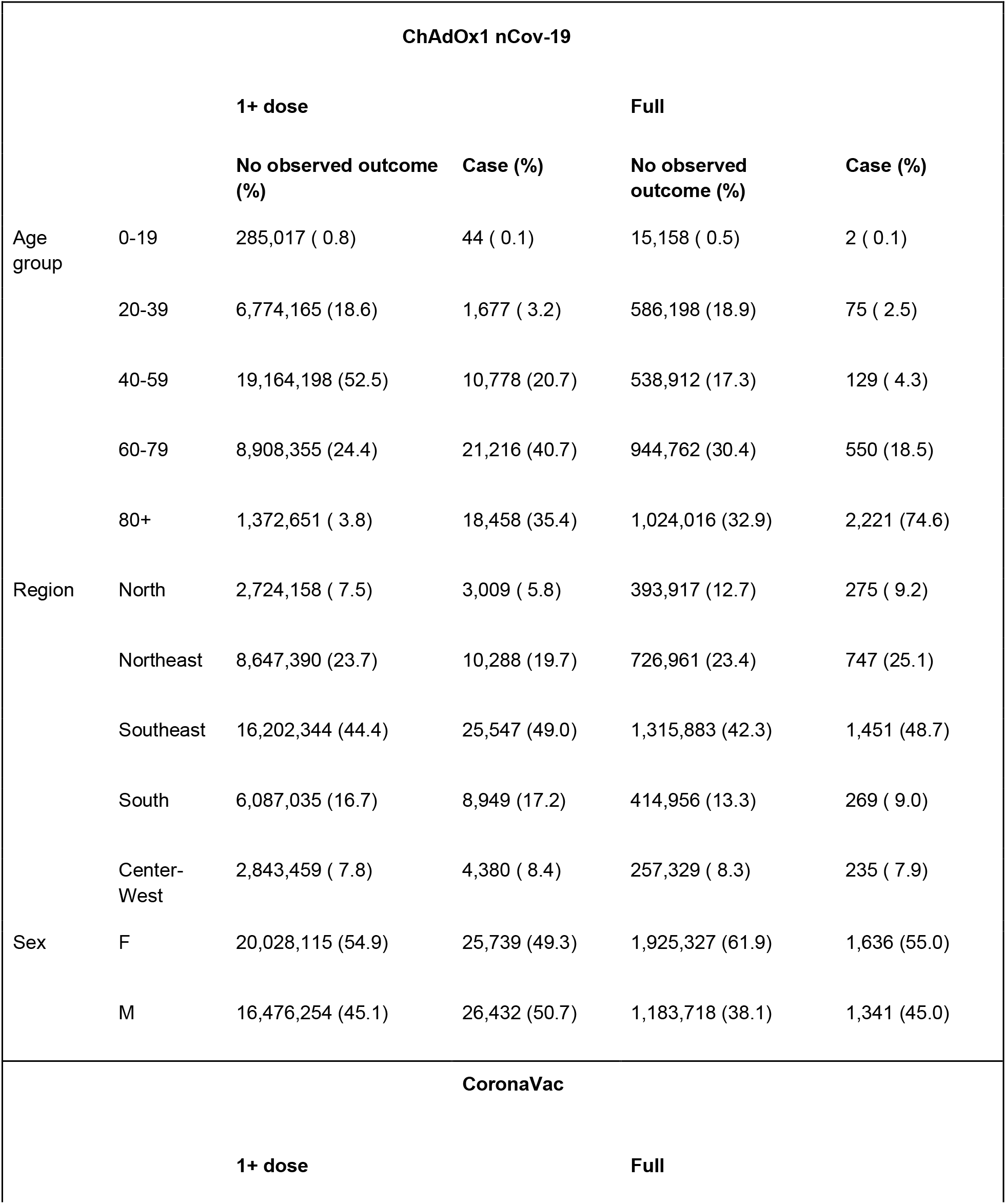
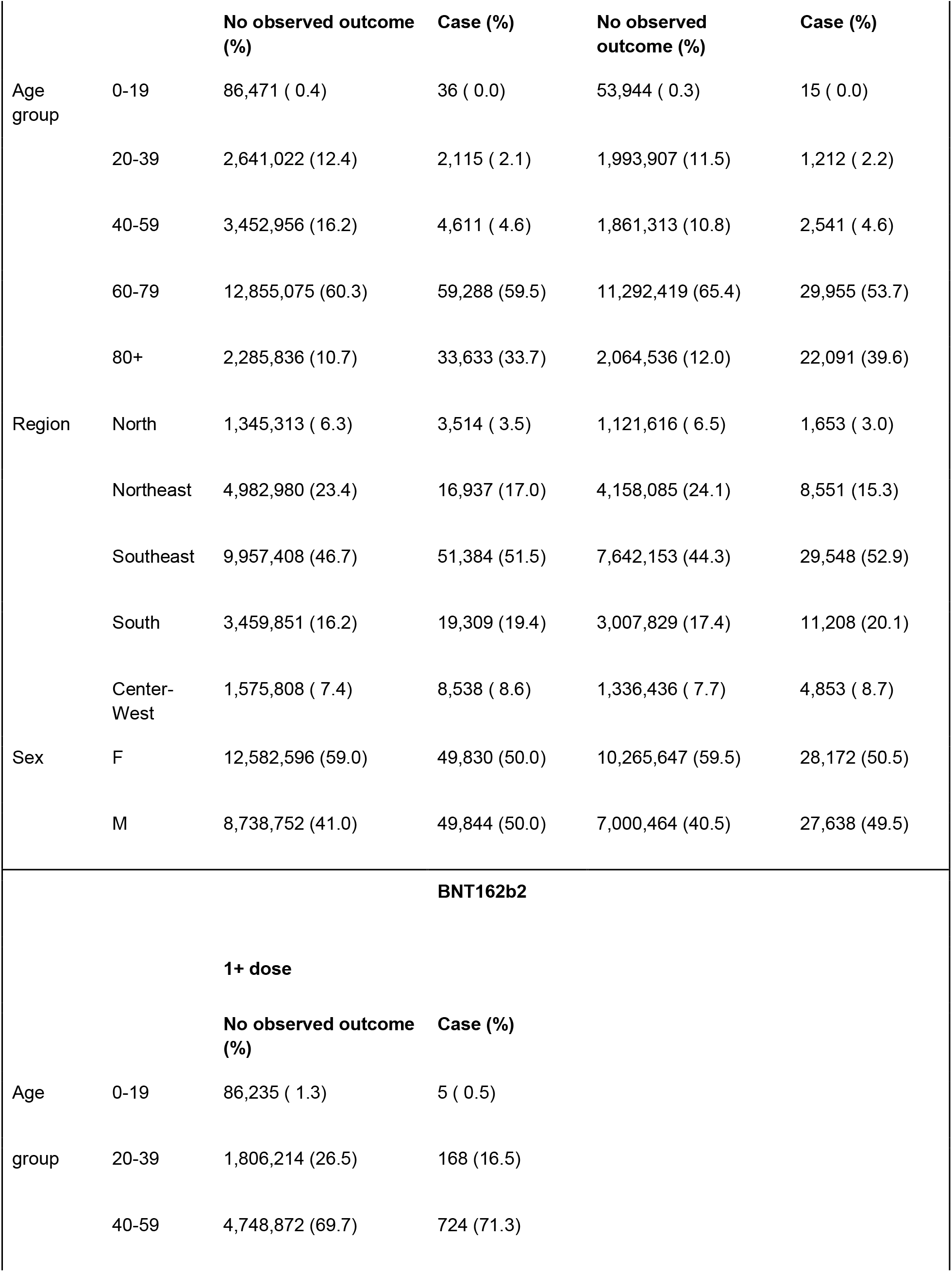
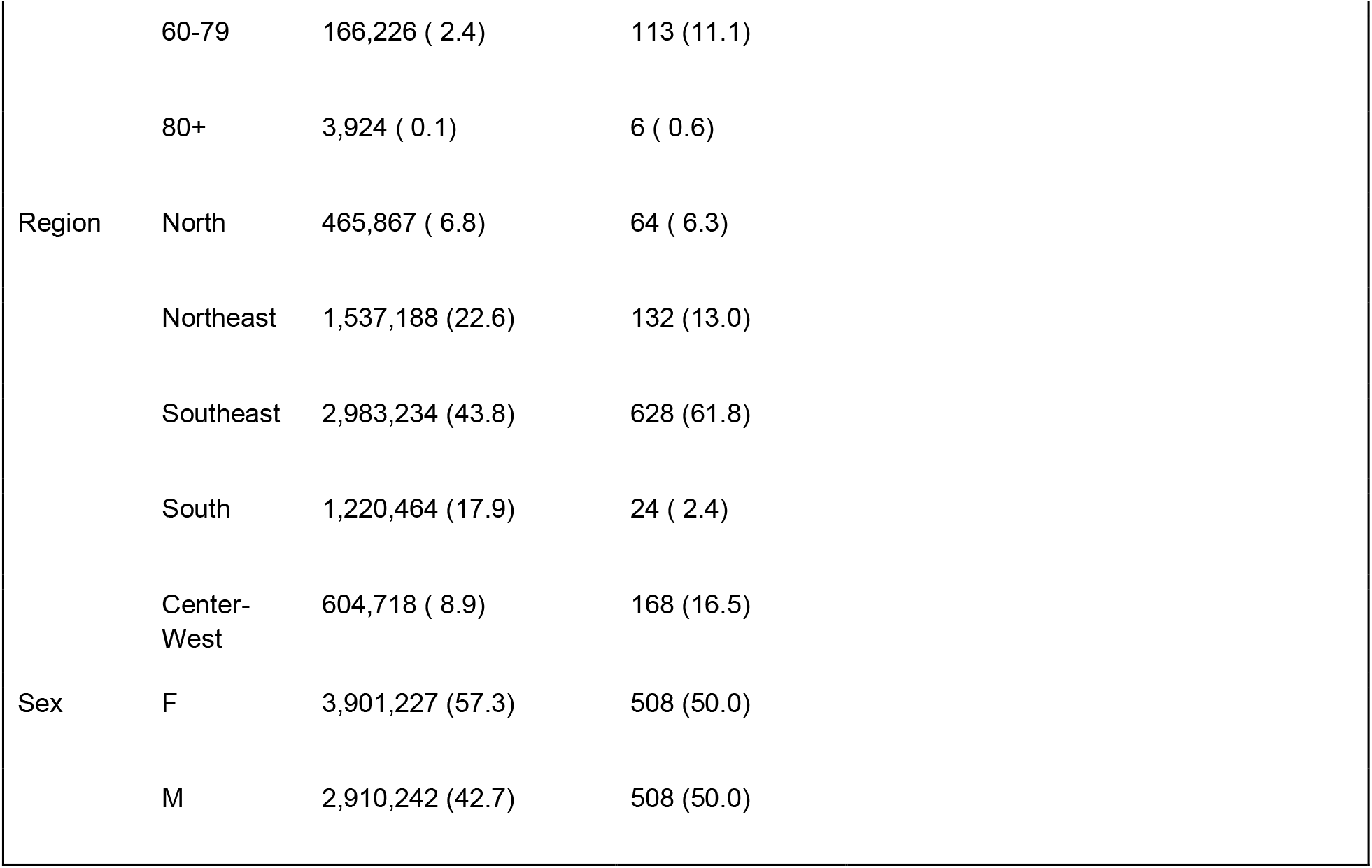
Severe COVID-19 cases among individuals in the cohort receiving ChAdOx1 nCov-19, CoronaVac, or BNT162b2 vaccines, by age group, region and sex, in Brazil from vaccination T0 to July 17, 2021. Values in percentages indicate the proportions of each stratum item by the total in the stratum/vaccine/regimen.

| ChAdOx1 nCov-19 |  |  |  |  |  |
| --- | --- | --- | --- | --- | --- |
|  |  | 1+ dose |  | Full |  |
|  |  | No observed outcome (%) | Case (%) | No observed outcome (%) | Case (%) |
| Age group | 0-19 | 285,017 ( 0.8) | 44 ( 0.1) | 15,158 ( 0.5) | 2 ( 0.1) |
|  | 20-39 | 6,774,165 (18.6) | 1,677 ( 3.2) | 586,198 (18.9) | 75 ( 2.5) |
|  | 40-59 | 19,164,198 (52.5) | 10,778 (20.7) | 538,912 (17.3) | 129 ( 4.3) |
|  | 60-79 | 8,908,355 (24.4) | 21,216 (40.7) | 944,762 (30.4) | 550 (18.5) |
|  | 80+ | 1,372,651 ( 3.8) | 18,458 (35.4) | 1,024,016 (32.9) | 2,221 (74.6) |
| Region | North | 2,724,158 ( 7.5) | 3,009 ( 5.8) | 393,917 (12.7) | 275 ( 9.2) |
|  | Northeast | 8,647,390 (23.7) | 10,288 (19.7) | 726,961 (23.4) | 747 (25.1) |
|  | Southeast | 16,202,344 (44.4) | 25,547 (49.0) | 1,315,883 (42.3) | 1,451 (48.7) |
|  | South | 6,087,035 (16.7) | 8,949 (17.2) | 414,956 (13.3) | 269 ( 9.0) |
|  | Center-West | 2,843,459 ( 7.8) | 4,380 ( 8.4) | 257,329 ( 8.3) | 235 ( 7.9) |
| Sex | F | 20,028,115 (54.9) | 25,739 (49.3) | 1,925,327 (61.9) | 1,636 (55.0) |
|  | M | 16,476,254 (45.1) | 26,432 (50.7) | 1,183,718 (38.1) | 1,341 (45.0) |
| CoronaVac |  |  |  |  |  |
|  |  | 1+ dose |  | Full |  |

|  |  | No observed outcome (%) | Case (%) | No observed outcome (%) | Case (%) |
| --- | --- | --- | --- | --- | --- |
| Age group | 0-19 | 86,471 ( 0.4) | 36 ( 0.0) | 53,944 ( 0.3) | 15 ( 0.0) |
|  | 20-39 | 2,641,022 (12.4) | 2,115 ( 2.1) | 1,993,907 (11.5) | 1,212 ( 2.2) |
|  | 40-59 | 3,452,956 (16.2) | 4,611 ( 4.6) | 1,861,313 (10.8) | 2,541 ( 4.6) |
|  | 60-79 | 12,855,075 (60.3) | 59,288 (59.5) | 11,292,419 (65.4) | 29,955 (53.7) |
|  | 80+ | 2,285,836 (10.7) | 33,633 (33.7) | 2,064,536 (12.0) | 22,091 (39.6) |
| Region | North | 1,345,313 ( 6.3) | 3,514 ( 3.5) | 1,121,616 ( 6.5) | 1,653 ( 3.0) |
|  | Northeast | 4,982,980 (23.4) | 16,937 (17.0) | 4,158,085 (24.1) | 8,551 (15.3) |
|  | Southeast | 9,957,408 (46.7) | 51,384 (51.5) | 7,642,153 (44.3) | 29,548 (52.9) |
|  | South | 3,459,851 (16.2) | 19,309 (19.4) | 3,007,829 (17.4) | 11,208 (20.1) |
|  | Center-West | 1,575,808 ( 7.4) | 8,538 ( 8.6) | 1,336,436 ( 7.7) | 4,853 ( 8.7) |
| Sex | F | 12,582,596 (59.0) | 49,830 (50.0) | 10,265,647 (59.5) | 28,172 (50.5) |
|  | M | 8,738,752 (41.0) | 49,844 (50.0) | 7,000,464 (40.5) | 27,638 (49.5) |
| **BNT162b2** |  |  |  |  |  |
| **1+ dose** |  |  |  |  |  |
|  |  | No observed outcome (%) | Case (%) |  |  |
| Age group | 0-19 | 86,235 ( 1.3) | 5 ( 0.5) |  |  |
|  | 20-39 | 1,806,214 (26.5) | 168 (16.5) |  |  |
|  | 40-59 | 4,748,872 (69.7) | 724 (71.3) |  |  |

|  |  |  |  |
| --- | --- | --- | --- |
|  | 60-79 | 166,226 ( 2.4) | 113 (11.1) |
|  | 80+ | 3,924 ( 0.1) | 6 ( 0.6) |
| Region | North | 465,867 ( 6.8) | 64 ( 6.3) |
|  | Northeast | 1,537,188 (22.6) | 132 (13.0) |
|  | Southeast | 2,983,234 (43.8) | 628 (61.8) |
|  | South | 1,220,464 (17.9) | 24 ( 2.4) |
|  | Center-West | 604,718 ( 8.9) | 168 (16.5) |
| Sex | F | 3,901,227 (57.3) | 508 (50.0) |
|  | M | 2,910,242 (42.7) | 508 (50.0) |

**Table 3:**
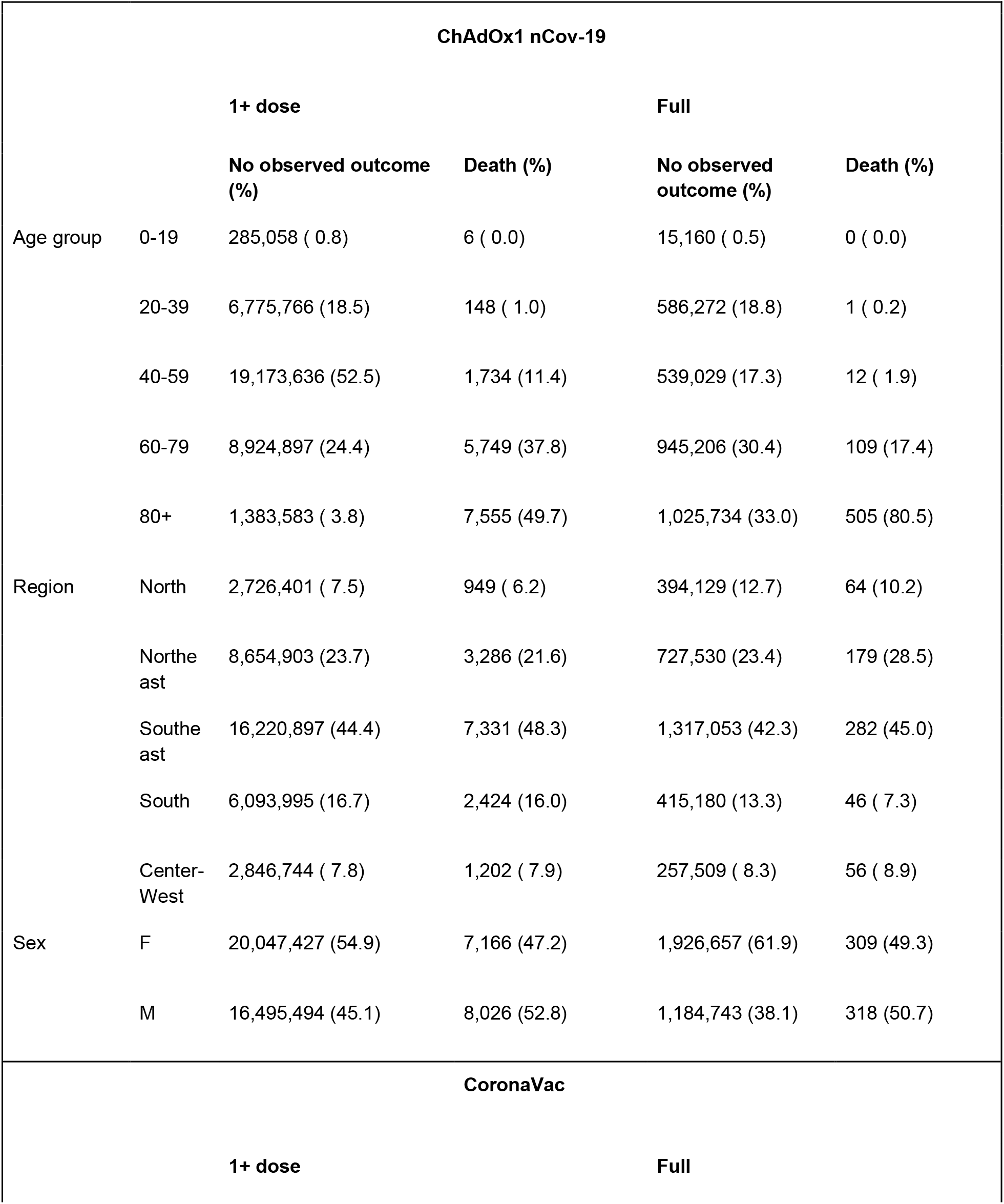
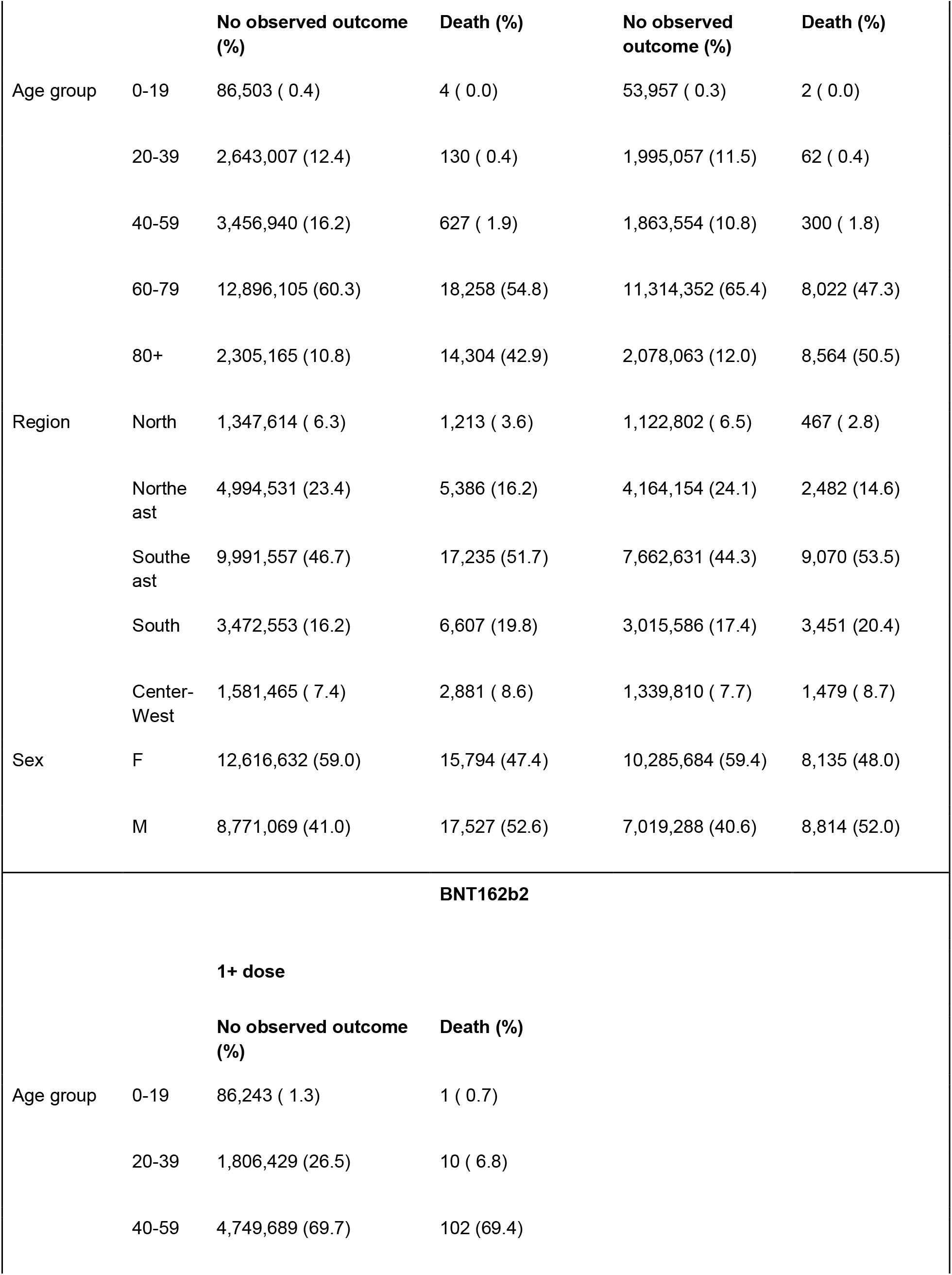
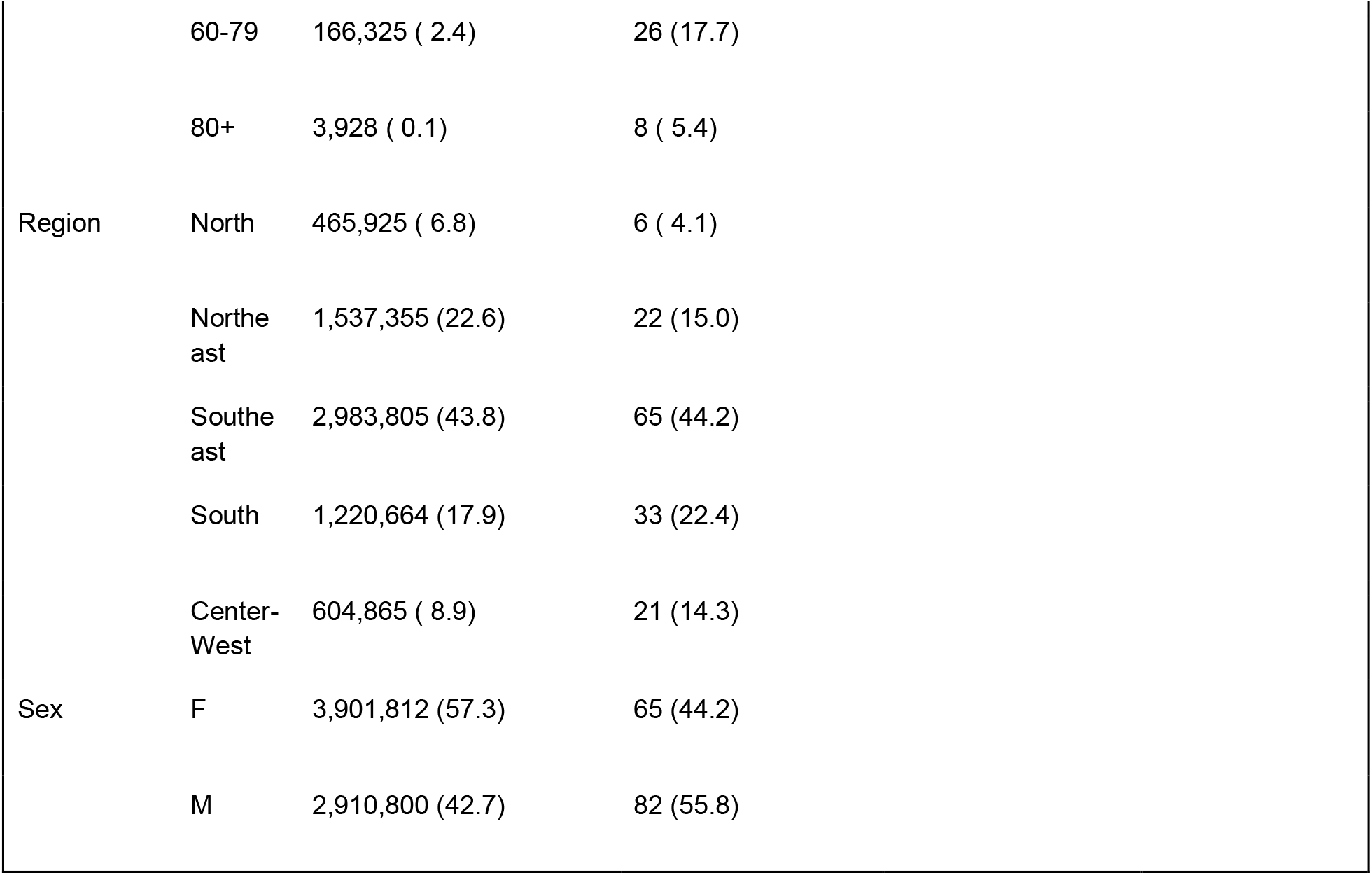
Deaths due to severe COVID-19 among individuals in the cohort receiving ChAdOx1 nCov-19, CoronaVac, and BNT162b2 vaccines, by age group, region and sex, Brazil from vaccination T0 to July 17, 2021. Values in percentages indicate the proportions of each stratum item by the total in the stratum/vaccine/regimen.

| ChAdOx1 nCov-19 |  |  |  |  |  |
| --- | --- | --- | --- | --- | --- |
|  |  | 1+ dose |  | Full |  |
|  |  | No observed outcome (%) | Death (%) | No observed outcome (%) | Death (%) |
| Age group | 0-19 | 285,058 ( 0.8) | 6 ( 0.0) | 15,160 ( 0.5) | 0 ( 0.0) |
|  | 20-39 | 6,775,766 (18.5) | 148 ( 1.0) | 586,272 (18.8) | 1 ( 0.2) |
|  | 40-59 | 19,173,636 (52.5) | 1,734 (11.4) | 539,029 (17.3) | 12 ( 1.9) |
|  | 60-79 | 8,924,897 (24.4) | 5,749 (37.8) | 945,206 (30.4) | 109 (17.4) |
|  | 80+ | 1,383,583 ( 3.8) | 7,555 (49.7) | 1,025,734 (33.0) | 505 (80.5) |
| Region | North | 2,726,401 ( 7.5) | 949 ( 6.2) | 394,129 (12.7) | 64 (10.2) |
|  | Northeast | 8,654,903 (23.7) | 3,286 (21.6) | 727,530 (23.4) | 179 (28.5) |
|  | Southeast | 16,220,897 (44.4) | 7,331 (48.3) | 1,317,053 (42.3) | 282 (45.0) |
|  | South | 6,093,995 (16.7) | 2,424 (16.0) | 415,180 (13.3) | 46 ( 7.3) |
|  | Center-West | 2,846,744 ( 7.8) | 1,202 ( 7.9) | 257,509 ( 8.3) | 56 ( 8.9) |
| Sex | F | 20,047,427 (54.9) | 7,166 (47.2) | 1,926,657 (61.9) | 309 (49.3) |
|  | M | 16,495,494 (45.1) | 8,026 (52.8) | 1,184,743 (38.1) | 318 (50.7) |
| CoronaVac |  |  |  |  |  |
|  |  | 1+ dose |  | Full |  |

|  |  | No observed outcome (%) | Death (%) | No observed outcome (%) | Death (%) |
| --- | --- | --- | --- | --- | --- |
| Age group | 0-19 | 86,503 ( 0.4) | 4 ( 0.0) | 53,957 ( 0.3) | 2 ( 0.0) |
|  | 20-39 | 2,643,007 (12.4) | 130 ( 0.4) | 1,995,057 (11.5) | 62 ( 0.4) |
|  | 40-59 | 3,456,940 (16.2) | 627 ( 1.9) | 1,863,554 (10.8) | 300 ( 1.8) |
|  | 60-79 | 12,896,105 (60.3) | 18,258 (54.8) | 11,314,352 (65.4) | 8,022 (47.3) |
|  | 80+ | 2,305,165 (10.8) | 14,304 (42.9) | 2,078,063 (12.0) | 8,564 (50.5) |
| Region | North | 1,347,614 ( 6.3) | 1,213 ( 3.6) | 1,122,802 ( 6.5) | 467 ( 2.8) |
|  | Northeast | 4,994,531 (23.4) | 5,386 (16.2) | 4,164,154 (24.1) | 2,482 (14.6) |
|  | Southeast | 9,991,557 (46.7) | 17,235 (51.7) | 7,662,631 (44.3) | 9,070 (53.5) |
|  | South | 3,472,553 (16.2) | 6,607 (19.8) | 3,015,586 (17.4) | 3,451 (20.4) |
|  | Center-West | 1,581,465 ( 7.4) | 2,881 ( 8.6) | 1,339,810 ( 7.7) | 1,479 ( 8.7) |
| Sex | F | 12,616,632 (59.0) | 15,794 (47.4) | 10,285,684 (59.4) | 8,135 (48.0) |
|  | M | 8,771,069 (41.0) | 17,527 (52.6) | 7,019,288 (40.6) | 8,814 (52.0) |
| **BNT162b2** |  |  |  |  |  |
|  |  | **1+ dose** |  |  |  |
|  |  | **No observed outcome (%)** | **Death (%)** |  |  |
| Age group | 0-19 | 86,243 ( 1.3) | 1 ( 0.7) |  |  |
|  | 20-39 | 1,806,429 (26.5) | 10 ( 6.8) |  |  |
|  | 40-59 | 4,749,689 (69.7) | 102 (69.4) |  |  |

|  |  |  |  |
| --- | --- | --- | --- |
|  | 60-79 | 166,325 ( 2.4) | 26 (17.7) |
|  | 80+ | 3,928 ( 0.1) | 8 ( 5.4) |
| Region | North | 465,925 ( 6.8) | 6 ( 4.1) |
|  | Northeast | 1,537,355 (22.6) | 22 (15.0) |
|  | Southeast | 2,983,805 (43.8) | 65 (44.2) |
|  | South | 1,220,664 (17.9) | 33 (22.4) |
|  | Center-West | 604,865 ( 8.9) | 21 (14.3) |
| Sex | F | 3,901,812 (57.3) | 65 (44.2) |
|  | M | 2,910,800 (42.7) | 82 (55.8) |

The overall effectiveness of the vaccination program independent of the vaccine and age was 55.3% (54.9 - 55.7%) preventing severe COVID-19 cases, considering the two-dose regimen. For ChAdOx1 nCov-19 and CoronaVac, effectiveness for severe cases was, respectively, 72.8% (95% CrI: 71.8 - 73.8%) and 53.4% (95% CrI: 53.0% - 53.8%), on a two-dose regimen, independent of age. Effectiveness preventing severe cases with full immunization considering all vaccines exceeded 50% in age groups of 20-39, 40-59, 60-79 years old, preventing severe cases, and all age groups for death outcomes (Table 4). Effectiveness of ChAdOx1 nCov-19 was highest preventing severe cases among individuals 40-59 years old, reaching 90.4% [95% Credibility Interval (CrI): 88.7-92.0%]. Effectiveness of CoronaVac was highest among individuals 40-59 years old reaching 71.0% (95% CrI: 69.8-72.1) for severe cases. Effectiveness of BNT162b2 with at least one dose in preventing severe cases was highest among individuals 40-59 years old, and 60-79 years old, reaching 81.2 (95% CrI: 79.9--82.4) and 81.6% (95% CrI: 78.3--84.6), respectively, in these groups. Most importantly, the estimates for preventing severe cases in the group 60-79 years old at 79.6% (95% CrI: 77.8-81.3), for ChAdOx1 nCov-19, and 60.4 (95% CrI: 59.9-60.9), for CoronaVac, decreased to 66.7% (95%CrI: 65.1-68.1) and 29.6% (95% CrI: 28.5-30.8), for ChAdOx1 nCov-19 and CoronaVac, respectively, with the group of 80+ years.

**Table 4:** Vaccination effectiveness for individuals with at least first dose and fully immunized. The label “All vaccines” refers to the overall evaluation of the vaccination program.

|  |  | At least first dose |  | Fully immunized |  |
| --- | --- | --- | --- | --- | --- |
| Vaccine | Age group | Severe cases/deaths | Deaths | Severe cases/deaths | Deaths |
|  |  | Est. (95% CrI) | Est. (95% CrI) | Est. (95% CrI) | Est. (95% CrI) |
| All vaccines |  |  |  |  |  |
|  | 20-39 | 54.9 (53.4--56.3) | 72.2 (68.8--75.3) | 61.7 (59.6--63.8) | 83.7 (79.5--87.5) |
|  | 40-59 | 66.8 (66.2--67.3) | 75.3 (74.3--76.3) | 73.6 (72.6--74.6) | 84.5 (82.8--86.2) |
|  | 60-79 | 55.1 (54.7--55.4) | 64.4 (63.9--64.9) | 61.0 (60.5--61.5) | 71.8 (71.2--72.4) |
|  | 80+ | 23.9 (22.9--24.9) | 35.3 (34.0--36.5) | 35.9 (34.9--36.9) | 51.4 (50.2--52.6) |
| CoronaVac |  |  |  |  |  |
|  | 20-39 | 48.5 (46.2--50.7) | 72.5 (67.5--77.1) | 58.4 (56--60.7) | 81.5 (76.6--85.8) |
|  | 40-59 | 65.1 (64.1--66.2) | 76.1 (74.2--77.9) | 71.0 (69.8--72.1) | 82.7 (80.7--84.6) |
|  | 60-79 | 50.2 (49.7--50.6) | 58.9 (58.2--59.5) | 60.4 (59.9--60.9) | 71.2 (70.6--71.9) |
|  | 80+ | 21.8 (20.7--23) | 33.2 (31.7--34.6) | 29.6 (28.5--30.8) | 45.0 (43.6--46.4) |
| ChAdOx1 nCov-19 |  |  |  |  |  |
|  | 20-39 | 59.4 (57.4--61.3) | 69.8 (64.6--74.5) | 83.7 (79.8--87.2) | 97.9 (93.5--99.8) |
|  | 40-59 | 65.0 (64.3--65.6) | 72.7 (71.4--74.0) | 90.4 (88.7--92.0) | 95.6 (92.7--97.8) |
|  | 60-79 | 63.9 (63.4--64.4) | 74.5 (73.8--75.2) | 79.6 (77.8--81.3) | 89.5 (87.4--91.4) |
|  | 80+ | 26.9 (25.6--28.3) | 38.4 (36.7--40.0) | 66.7 (65.1--68.1) | 84.6 (83.3--85.9) |
| BNT162b2* |  |  |  |  |  |
|  | 20-39 | 64.7 (59.8--69.3) | 86.1 (76.9--93.8) | - | - |
|  | 40-59 | 81.2 (79.9--82.4) | 89.9 (87.8--91.8) | - | - |
|  | 60-79 | 81.6 (78.3--84.6) | 89.6 (85.1--93.2) | - | - |
|  | 80+ | 33.0 (-10.7--65.1) | 8.6 (-67.9--59.6) | - | - |

\* Not evaluated in fully immunized

Vaccine effectiveness was higher when analyzing death outcomes for all three vaccines. For CoronaVac fully immunized individuals, effectiveness was highest at 82.7% (95% CrI: 80.7-84.6%) in the group 40-59 years old. For full immunization with ChAdOx1 nCov-19 vaccine, the effectiveness preventing deaths was highest at 97.9% (95% CrI: 93.5--99.8) among 20-39 year old individuals and higher than 80% in all age groups. BNT162b2 effectiveness in the Brazilian population groups, which included partially immunized individuals, was highest in groups 40-59 and 60-79, respectively, estimated at 89.9% (95% CrI: 87.8--91.8) and 89.6 (95% CrI: 85.1-- 93.2) on preventing death. Effectiveness preventing deaths diminished with age, most significantly in the group of 80+ years old.

The effectiveness estimates on full regimen in all Brazilian regions were higher in the group 60-79 years old than those obtained in the group of 80+, both for CoronaVac and ChAdOx1 nCov-19, with severe cases and deaths as outcomes (Figure S2). The difference between effectiveness estimates in both groups is beyond the statistically significant bands for CoronaVac, except for severe cases in the North region. In the South and Southeast regions, the lowest estimates are below 50%. The differences between the effectiveness of ChAdOx1 nCov-19 vaccination in these groups were more significant in analyzing severe cases. Figure S3 shows the trends when considering the overall vaccination program.

Figure S4 shows how the incidence by age groups of severe COVID-19 cases peaked in March 2021 in macroregions of Brazil. Thereafter, incidences have declined in all age groups, most notably in 60-79. The 80+ year-old group, however, remained with the highest incidence, which plateaued in the Center-West, South, and Southeast regions. The great majority of sequenced COVID-19 cases from this study period in all regions were attributed to the gamma VOC (Figure S5).

## Discussion

The effectiveness of the Brazilian vaccination program against COVID-19, including the effectiveness of the deployed vaccines, ChAdOx1 nCov-19, CoronaVac, and BNT162b2, up to mid-July, varied considerably with age in this massive data analysis of severe COVID-19 cases and death as outcomes of interest. Results from this work were shared with the National Immunization Program and discussed within the National Advisory Committee on Immunization.

The effectiveness estimates reached high values for severe cases in most age groups, even higher figures preventing death outcomes and tight uncertainty intervals. In the group of non-elderly adults (20-39, 40-59), vaccine effectiveness for CoronaVac fully-vaccinated individuals went over 80% preventing death and over 70% preventing severe disease among 40-59 individuals. CoronaVac was the most frequent vaccine among fully-vaccinated individuals, hence putting the numbers of the overall vaccination program closer to the CoronaVac estimates. Effectiveness preventing severe disease was above 80% for the 20-39, 40-59, 60-79 age groups, considering partially or fully vaccinated with BNT162b2. Effectiveness estimates in individuals receiving ChAdOx1 nCov-19 full immunization exceeded 80% for preventing severe disease in non-elderly groups, and 60% for the elderly groups, and were higher than 90% for preventing death in non-elderly and 80% in elderly groups.

Serious concerns remain due to the diminishing effectiveness observed as age increases, particularly differences between 60-79 and 80+ age groups. Immunosenescence, a potentially limited duration of immunity, and earlier immunization in the 80+ group could also have led to lower effectiveness, given that this group had priority to start the immunization, in accordance with some studies (5,6). The states in the South and Southeast regions are the most populated and concentrated most of this difference. Conversely, the states in the North region had less reduction. Some of these states already had a high circulation of the gamma variant when vaccination started, with a higher incidence of severe COVID-19 cases during the first months, whereas both an increase of severe COVID-19 incidence and gamma variant predominance were observed a few weeks later in the other regions (Figures 3 and S3).

ChAdOx1 nCov-19 and CoronaVac were the most used vaccines in the period since most records of first dose were from ChAdOx1 nCov-19 receivers, and most people completing the immunization regimen were CoronaVac receivers. In addition, the number of vaccinated records in the age groups 40-59, 60-79, and 80+ indicate a significant but increasing immunization coverage, whereas numbers were relatively lower in the group 20-39. Therefore, our analyses were more robust for the former age groups.

The vaccination program likely played a major role in reducing severe cases and deaths over the weeks, especially after April, when vaccination coverage scaled up in several age groups. Accordingly, an early assessment via a demographic approach by Victora *et al*. showed changing COVID-mortality trends in Brazil with the increase of vaccination rates in the country (7). In Figure 3, while incidence kept clearly decreasing among 60-79 year old individuals during the period of late March to July in all regions, in which full vaccination coverage for that group was increasing, for the mostly unvaccinated or partially vaccinated age groups 20-39 and 40-59 there was a clear rebound between late April and early July. For those younger adults, this rebound reached values as high as those observed in March. A similar trend happened initially for the age group 80+, although by late April, cases stopped decreasing and reached a plateau, probably a consequence of significant vaccination coverage, combined with a sustained viral transmission in the population. Despite the overall decrease in incidence, the incidence in the 80+ year old group remained the highest, suggesting an effect of the lower vaccine’s effectiveness in this age group.

The methodology here has advantages over the Screening method, described in WHO guidelines (8), due to the use of rate ratios and stratification by age groups and states, a sound approach given the variability of incidences across a large country such as Brazil. In addition, it is similar to evaluation methods of other vaccination programs such as Influenza (9).

Effectiveness estimates of ChAdOx1 nCov-19 in Brazil are compatible with prior findings on its efficacy and prior effectiveness. Results in blind, randomized, controlled trials of ChAdOx1 nCov-19 indicated the efficacy of 62% in individuals receiving two regular doses, and 90% with combination of low dose/regular dose, with overall efficacy assessed at 70.4% (54.8 - 80.6%) (10), and subsequently assessed at 76·0% (59·3–85·9) after a first single dose and 81·3% (95% CI 60·3–91·2) (11). A test-negative case-control study to evaluate effectiveness of both BNT162b2 and ChAdOx1 nCov-19 found 73% (27% to 90%) for ChAdOx1 nCov-19 with a further reduction of 37% risk of hospitalization (12). Here, the effectiveness of at-least partially immunized individuals also included fully immunized individuals, however, in the case of ChAdOx1 nCov-19, the majority was still in a single-dose.

Efficacy of CoronaVac in a double-blind, randomized, controlled trial was estimated at 83·5% (65·4–92·1) after the second dose (13), whereas effectiveness in Chile indicated 87.5% (95% CI, 86.7 to 88.2) for the prevention of hospitalization, and 86.3% (95% CI, 84.5 to 87.9) for the prevention of Covid-19–related death (14). The same study indicated effectiveness of CoronaVac full immunization over 60 years of age at 85.3% (84.3–86.3) for hospitalization and 86.5% (84.6–88.1) for death. Thus, effectiveness in Brazil preventing severe cases was lower after a full CoronaVac regimen in the age groups 60-79 years old and over 80 years. Still, it was close to recent estimates of a test-negative case-control study that evaluated effectiveness over 70 years of age in São Paulo (15) and among healthcare workers in Manaus (16).

Efficacy of the BNT162b2 vaccine in randomized controlled trials was assessed at 95.0% (90.0– 97.9) (17). Effectiveness of BNT162b2 vaccination in Israel, evaluated in a case-control design, was at 87 % (55–100%) for hospitalized cases after seven days from regimen completion (18). Bernal e*t al*. found the effectiveness of BNT162b2 was 61% (51 - 69%) for symptomatic cases in a test-negative case-control design with a further reduction of 43% risk of hospitalization (12). In our study, the effectiveness of BNT162b2 was in general below these levels, yet evaluation of the BNT162b2 vaccine was more limited due to the smaller number of immunized individuals and shorter follow up time, especially for the elderly.

The impact of VOCs on vaccine effectiveness is an evolving issue. Few studies have evaluated the impact on humoral immune responses, with a modest reduction of responses in either ChAdOx1 nCov-19 or BNT162b2 in the presence of the delta variant, although with a more significant difference with a single dose, i.e., before the second dose (19), and lower levels of neutralizing antibodies in healthcare workers with a two-dose regimen of CoronaVac in Thailand for the alpha, beta, and delta VOC, with a more pronounced reduction for the delta VOC (20). Shapiro et al. provide a comprehensive review on the effectiveness of COVID-19 vaccines (21). Our methodology can be used to monitor vaccine effectiveness over time and will be applied to monitor the potential impact of these VOCs on vaccine effectiveness in Brazil.

A few factors related to confounding effects and time-varying exposure to SARS-CoV-2 may hinder the effectiveness evaluation. The analysis is sensitive to COVID-19 incidence over time, the occurrence of VOCs, and other interventions during the study period, such as restrictive measures in some municipalities and/or states. The easing of non-pharmacological measures after vaccination, such as less frequent use of masks and increased face-to-face social interactions without proper distancing and ventilation care, can induce a greater risk of infection. Prevalences of comorbidities and exposure risk can be confounders, particularly in the young adults, among which individuals with comorbidities and healthcare workers were initially prioritized for vaccination, inducing a potential selection bias. Such bias is likely less significant for individuals over 60 years old since vaccination in this group was mainly targeted by age. Furthermore, the methodology depends on the quality of notifications of vaccinating records and cases in databases and on projections of the resident population by age group in the states, since the most recent census data in Brazil is from 2010. Still, the methodology can be adapted to changing epidemiological scenarios, and its use on permanent monitoring of the effectiveness should be pursued to investigate potential confounders further.

Finally, permanent efforts are necessary to estimate the vaccination effectiveness in the various epidemiological contexts of Brazil and other countries. Results from a timely evaluation of vaccination effectiveness can be updated periodically and integrate regular surveillance, hence they are instrumental to be shared with authorities, in this case, the Brazilian Ministry of Health. Furthermore, considering the biological plausibility of immunosenescence and the possibility of decreased immunity over time, especially in the context of VOC spreading, more studies with the elderly are needed towards potential revaccination to avoid serious COVID-19 illness and deaths.

## Data Availability

Data in this study comes from a database maintained by the Brazilian Ministry of Health (MoH), which provided the anonymized database for this analyses.

## Acknowledgement

Authors would like to thank GISAID for the access to the database of lineages of SARS-CoV-2 virus in Brazil and Ana Tereza R. de Vasconcelos and Luiz G. P. de Almeida for helping with obtaining the GISAID information from Brazil. The authors had support from INOVA Fiocruz, CAPES, CNPq (Refs. 441057/2020-9, 309569/2019-2). The Brazilian Ministry of Health supported the project and provided the anonymized database.

## Data Statement

Data in this study comes from a database maintained by the Brazilian Ministry of Health (MoH), which provided the anonymized database for the analyses.

## Ethics Statement

The study was conducted in accordance with fundamental ethical principles of the Declaration of Helsinki and the Brazilian National Health Council on research involving human beings. The study protocol was approved by the Research Ethics Committee of the Evandro Chagas National Institute of Infectious Diseases-Fiocruz (CAAE: 51567721.9.0000.5262).

