## Supplementary Material for "Effectiveness of Mass Vaccination in Brazil against Severe COVID-19 Cases"

Daniel A.M. Villela[1,*], Tatiana Guimarães de Noronha[2,3], Leonardo S. Bastos[1], Antonio G. Pacheco[1], Oswaldo G Cruz[1], Luiz Max Carvalho[4], Claudia Torres Codeço[1], Marcelo Ferreira da Costa Gomes[1], Flávio Codeço Coelho[4,5], Laís Picinini Freitas[1], Raquel Martins Lana[1], Victor Bertollo Gomes Porto[6], Luiz Antônio Bastos Camacho[7], Claudio José Struchiner[4,8]

1. Program of Scientific Computing, Fundação Oswaldo Cruz, Brazil
2. Institute of Technology for Immunobiologicals, Fundação Oswaldo Cruz, Brazil
3. School of Medicine, Universidade Federal Fluminense, Brazil
4. School of Applied Mathematics - Fundação Getúlio Vargas, Brazil
5. Institute of Global Health - University of Geneva, Switzerland.
6. National Immunization Program, Ministry of Health, Brazil
7. National School of Public Health, Fundação Oswaldo Cruz, Brazil
8. Institute of Social Medicine, Universidade do Estado do Rio de Janeiro, Brazil

### Methods

#### Study design

Semi-ecological study of a national health-record cohort with more than 66.3 million records, including over 1 million severe COVID-19 cases, where individual data were evaluated for tracking both the outcomes of interest (severe COVID-19 cases and deaths) and their vaccination status over time.

### Data Linkage

Records were linked sequentially by the Individual Taxpayer Registration (CPF) and the combination of full name and full mother's name. The remaining records were then linked probabilistically by a concatenated variable including the full name and full mother's name using a blocking strategy, which used a variable given by the concatenated string including the code of the municipality of residence, the individual age, gender, the first two letters of the individual's name, and the mother's name. This variable allowed efficiency and validation of the probabilistic linking algorithm. Records with a Levenshtein distance of 4 or less were considered a match. This threshold was chosen after a validation step. The linkage was performed by the Ministry of Health itself, which provided the resulting dataset already linked and anonymized to the researchers involved in this study. Authors not affiliated with the Ministry of Health did not have access to nominal data from the immunization program at any time.

### Statistical modeling and analysis

Cases of severe COVID-19 were aggregated by age group (20 - 39, 40 - 59, 60 - 79, and 80+ years old), vaccination status (vaccinated/unvaccinated), and the states of residency. In the model, given a time window from T0 to a time $t$, a total of $Y_i$ cases are observed, where $i$ represents a combination of age group, state and vaccination status. The age group $a(i)$ and the pair $h(i)$ of age group and state are necessary for comparing the risks of occurrence of one of the outcomes. Also, for each instance of the vaccination status $v_i$ ($v_i$=1, vaccinated; $v_i$=0, unvaccinated) it is important to distinguish the risk between vaccinated and unvaccinated individuals. For each individual in the cohort, there are periods $\tau_u$, and $\tau_v$ when the individual has unvaccinated and vaccinated status, defined by the date of vaccination plus 14 days. The number $D_i$ denotes the total person-time contributed by individuals to each combination of age group, geographic region and vaccination status, entering the model as an offset. In the case of vaccinated individuals, the person-time contribution is the sum of all vaccinated time $\tau_v$ over all individuals in the combination. The proportion $c_i$ gives the coverage of vaccination over persons and time, measured by the ratio between contribution of vaccinated person-time and the total person-time in the study. For non-vaccination combinations, the person-time component $D_i$ is given by the fraction $P_i$ $(1-c_i)$ $t_i$, where $P_i$ is the population estimate in the age group $a(i)$, and $t_i$ is the time from T0+14 to time $t$, and in the case of BNT162b2 vaccination, from T103+14 to t, due to a later start.

Cases are described by a mixed-effects Poisson model:

$Yi \sim Poisson(\lambda_i)$,

where $\log(\lambda_i) = \log(D_i) + \gamma_{h(i)} + \beta_{a(i)} v_i$, $\gamma_{h(i)}$ and $\beta_{a(i)}$ are random effects, in particular $\beta_{a(i)}$ is an age-varying effect.

A Bayesian analysis allows estimation of all coefficients, including $\beta_{a(i)}$ and the parameters $\gamma_{h(i)}$, and other transformed quantities. This estimation permits to obtain the rate ratio between the rate $R_v$ of severe COVID-19 event for a vaccinated group and the rate $R_u$ for an unvaccinated group, given by $R_v/R_u = exp(\beta_{a(i)})$. Vaccine effectiveness is given by 1 - $R_v/R_u$.

Without loss of generality, the same framework is used to analyze death as an outcome of interest. Separate analyses were performed considering the vaccination status to be at least partially immunized (1+ dose) and the fully immunized case. Also, the analysis varied from comprehensive covering the whole country, i.e. with all states, or by macroregions (North, Northeast, Center-West, Southeast, and South), i.e., focused on covering the states of the specific macroregion.

R platform (version 4.1.0) was used for data manipulation and exploratory data analysis and JAGS was used in the Bayesian setting for estimations with MCMC simulations.

**Results**

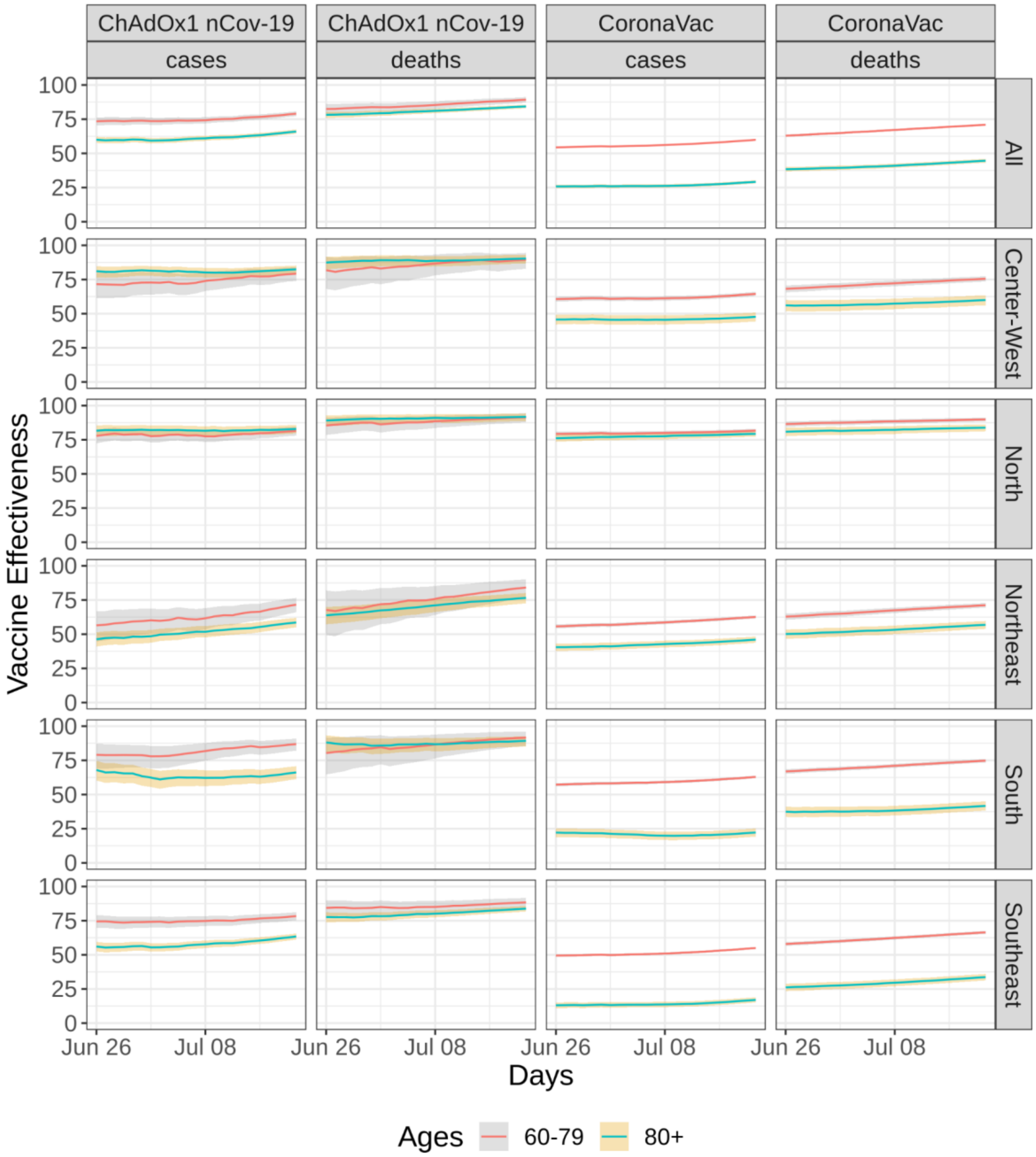

**Figure S1**. Evolution of effectiveness on full regimen over time for both CoronaVac and ChAdOx1 nCov-19 across regions of Brazil analyzing outcomes: severe COVID-19 cases and deaths. The mixed effects Poisson model was applied to a subset of the cohort database considering only information in the end of June to Mid-July, the last 24 days in the database. Lines indicate mean value estimates and color-shaded areas indicate 95% credibility intervals. The levels of effectiveness achieved by mid-July for the elderly groups across all regions shows a slightly increasing trend in the North and Northeast regions, likely due to ongoing vaccination rollout in these age groups.

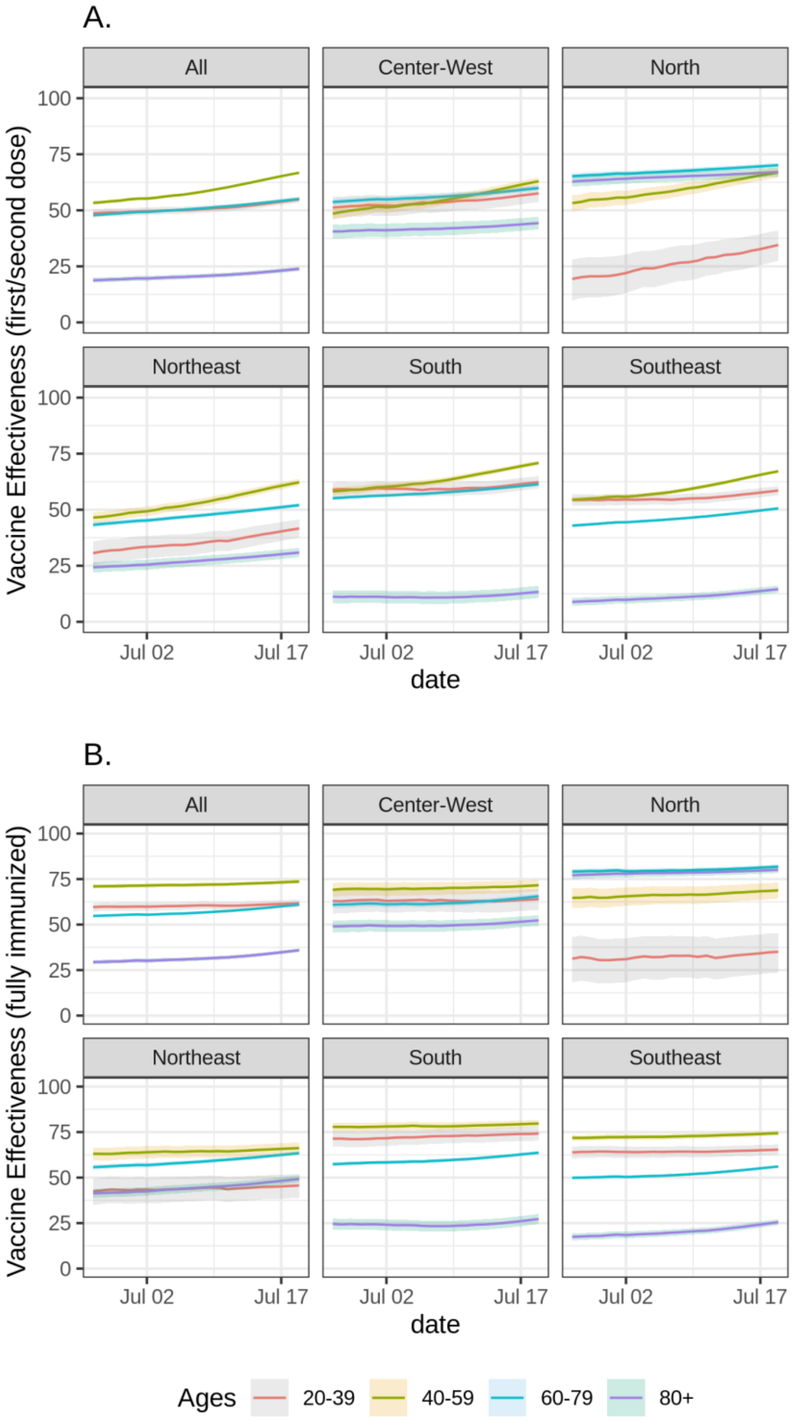

**Figure S2.** Effectiveness of the overall vaccination program over Brazilian regions when analyzing severe cases in the last 24 days of the study with (A) at least partially immunized individuals and (B) fully immunized individuals. Lines indicate mean value estimates and color-shaded areas indicate 95% credibility intervals.

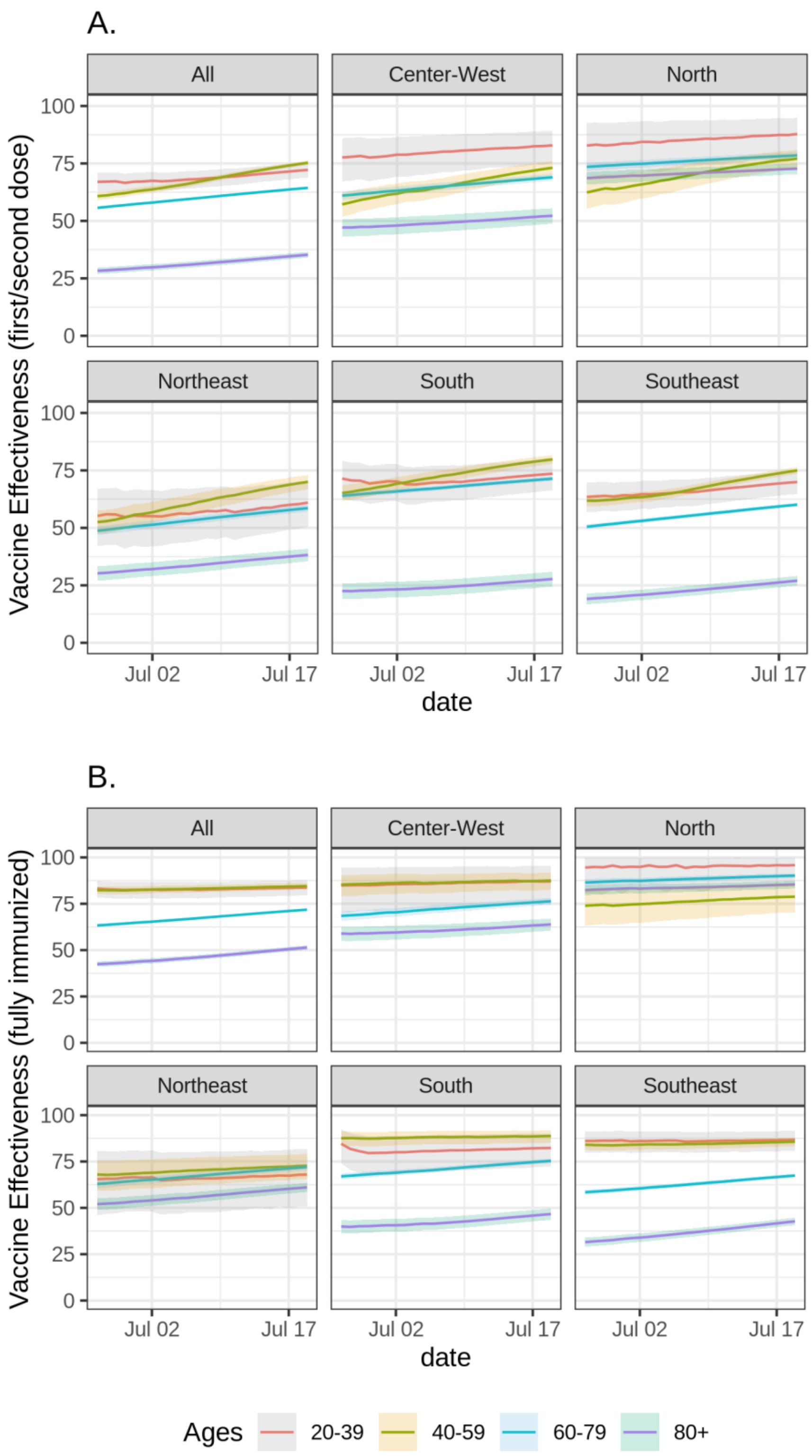

**Figure S3.** Effectiveness of the overall vaccination program over Brazilian regions when analyzing death only as outcomes in the last 24 days of the study with (A) at least partially immunized individuals and (B) fully immunized individuals. Lines indicate mean value estimates and color-shaded areas indicate 95% credibility intervals.

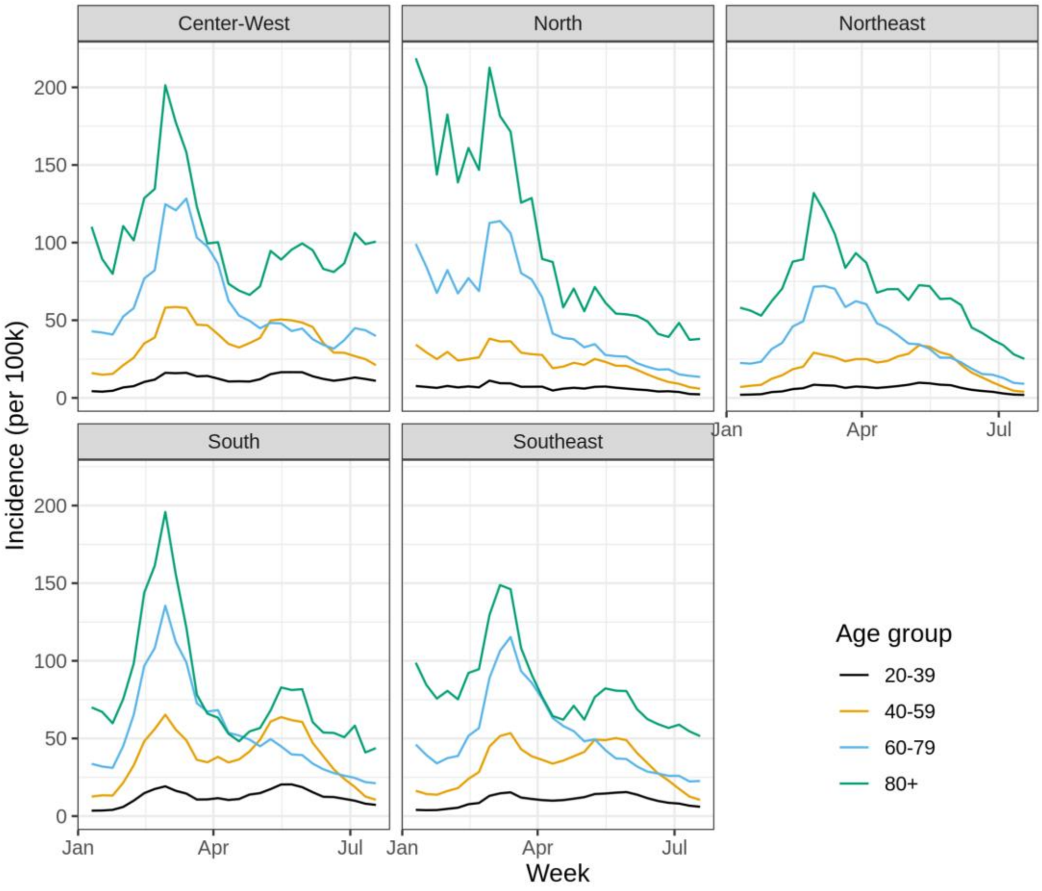

**Figure S4**. Weekly incidence of SARI by age groups in the five Brazilian macroregions during 2021. Values are shown in cases by 100 thousand in each age group. All records were collected from SIVEP in epidemiological week 33, but only showing until mid-July which is the maximum time frame in the effectiveness evaluation.

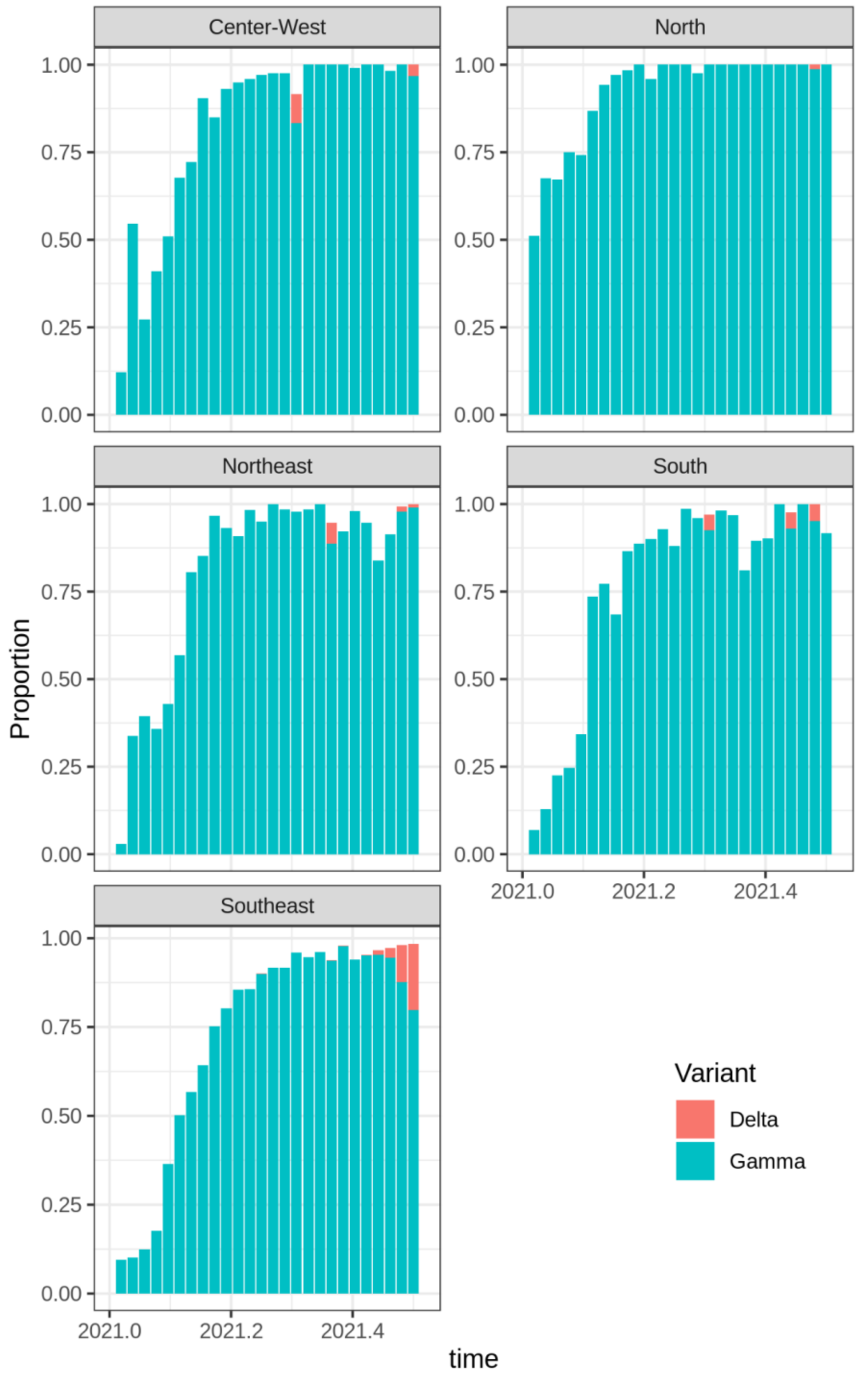

**Figure S5.** Frequency of delta and gamma variants of SARS-CoV-2 in all regions of Brazil in scale 0.0-1.0 during 2021.
